# Mathematical modeling of the SARS-CoV-2 epidemic in Qatar and its impact on the national response to COVID-19

**DOI:** 10.1101/2020.11.08.20184663

**Authors:** Houssein H. Ayoub, Hiam Chemaitelly, Shaheen Seedat, Monia Makhoul, Zaina Al Kanaani, Abdullatif Al Khal, Einas Al Kuwari, Adeel A. Butt, Peter Coyle, Andrew Jeremijenko, Anvar Hassan Kaleeckal, Ali Nizar Latif, Riyazuddin Mohammad Shaik, Hadi M. Yassine, Mohamed G. Al Kuwari, Hamad Eid Al Romaihi, Mohamed H. Al-Thani, Roberto Bertollini, Laith J. Abu Raddad

**Author notes:** Reprints or correspondence: Dr. Houssein H. Ayoub, Department of Mathematics, Statistics, and Physics, Qatar University, P.O. Box 2713, Doha, Qatar., Professor Laith J. Abu-Raddad, Infectious Disease Epidemiology Group, World Health Organization Collaborating Centre for Disease Epidemiology Analytics on HIV/AIDS, Sexually Transmitted Infections, and Viral Hepatitis, Weill Cornell Medicine - Qatar, Qatar Foundation - Education City, P.O.Box 24144, Doha, Qatar. **Funding:** The Biomedical Research Program and the Biostatistics, Epidemiology and Biomathematics Research Core at Weill Cornell Medicine-Qatar, Ministry of Public Health, and Hamad Medical Corporation. **Disclose funding received for this work:** others.

## Abstract

**Background:** Mathematical modeling constitutes an important tool for planning robust responses to epidemics. This study was conducted to guide the Qatari national response to the severe acute respiratory syndrome coronavirus 2 (SARS-CoV-2) epidemic. The study investigated the time course of the epidemic, forecasted healthcare needs, predicted the impact of social and physical distancing restrictions, and rationalized and justified easing of restrictions.

**Methods:** An age-structured deterministic model was constructed to describe SARS-CoV-2 transmission dynamics and disease progression throughout the population.

**Results:** The enforced social and physical distancing interventions flattened the epidemic curve, reducing the peaks for incidence, prevalence, acute-care hospitalization, and intensive care unit (ICU) hospitalizations by 87%, 86%, 76%, and 78%, respectively. The daily number of new infections was predicted to peak at 12,750 on May 23, and active-infection prevalence was predicted to peak at 3.2% on May 25. Daily acute-care and ICU-care hospital admissions and occupancy were forecast accurately and precisely. By October 15, 2020, the basic reproduction number *R*_0_ had varied between 1.07-2.78, and 50.8% of the population were estimated to have been infected (1.43 million infections). The proportion of actual infections diagnosed was estimated at 11.6%. Applying the concept of *R*_*t*_ tuning, gradual easing of restrictions was rationalized and justified to start on June 15, 2020, when *R*_*t*_ declined to 0.7, to buffer the increased interpersonal contact with easing of restrictions and to minimize the risk of a second wave. No second wave has materialized as of October 15, 2020, five months after the epidemic peak.

**Conclusions:** Use of modeling and forecasting to guide the national response proved to be a successful strategy, reducing the toll of the epidemic to a manageable level for the healthcare system.

## Introduction

Mathematical modeling has become a fundamental tool to guide surveillance of infectious diseases and emergency responses to epidemics [1-3]. Powered by surveillance and outbreak data, infection transmission models help monitor and predict epidemiological trends using real-time estimation of key indicators, such as incidence of infection, severe and critical disease cases, disease mortality, and basic reproduction number (*R*_0_; the number of secondary infections each infection would generate in a fully susceptible population [4]) [3].

Qatar is a peninsula located in the Arabian Gulf, with a diverse population of 2.8 million people [5]. Like other countries, Qatar has been affected by the severe acute respiratory syndrome coronavirus 2 (SARS-CoV-2) pandemic [6-9]. Yet, the nation mounted an evidence-informed national response, in which in addition to early case identification, isolation, and quarantine through contact tracing, diverse standardized and centralized sources of data were generated, including population-based surveys. This wealth of data provided a special opportunity to understand infection transmission dynamics, predict healthcare needs associated with the resulting disease, coronavirus disease 2019 (COVID-19) [10], and to inform the global epidemiology of this infection.

Qatar has a unique socio-demography that affected the transmission patterns of SARS-CoV-2 [8,11], a respiratory infection that propagates through social networks. Nearly 90% of the population are expatriates [5,12,13] with craft and manual workers (CMWs) constituting 60% of the population [14]. Of the national subpopulations, Indians (28%) constitute the largest population segment, followed by Bangladeshis (13%), Nepalese (13%), Qataris (11%), Egyptians (9%), and Filipinos (7%) [13]. The CMW population is predominantly male, single, and young, with the top three countries of origin being India, Bangladesh, and Nepal [14]. Most CMWs live in shared housing accommodations akin to dormitories [15].

This study was conducted to describe SARS-CoV-2 transmission dynamics in Qatar and to craft a national response using mathematical modeling of the epidemic time-course, predicting the impact of social and physical distancing restrictions and the impact of easing those restrictions, and forecasting healthcare needs, in terms of hospitalizations requiring acute-care and intensive care unit (ICU) beds. The study was initiated before the identification of the first laboratory-confirmed case of community transmission on March 6, 2020, and has continued to provide real-time projections and forecasts since then.

The overarching aim of the present article was to provide the technical tools and a “case study” to demonstrate how individual countries can use mathematical modeling to effectively craft national public-health responses and to formulate evidence-based policy decisions that minimize the epidemic’s toll on morbidity, mortality, societies, and economies.

## Methods

### Mathematical model

Building on our previously developed models [8,16-19], an age-structured, meta-population, deterministic mathematical model was constructed to describe SARS-CoV-2 transmission dynamics and disease progression (Figure S1 of Supplementary Material (SM)). The model stratified the Qatari population into groups (“compartments”) according to the major nationality groups (Indians, Bangladeshis, Nepalese, Qataris, Egyptians, Filipinos, and all other nationalities), age group by decile, infection status (infected, uninfected), severity of illness (asymptomatic/mild, severe, critical), and disease/hospitalization stage (severe, critical), using sets of coupled, nonlinear, differential equations. A detailed description of the model is available in the SM.

The risk of acquiring the infection varied between susceptible populations based on nationality, infectious contact rate per day, age-specific exposure/susceptibility to the infection, and subpopulation-mixing and age-mixing matrices parametrizing the mixing between individuals in different nationality and age groups. Following a latency period, infected individuals in the model develop an asymptomatic/mild, severe, or critical infection. The age-dependence of the proportions of infected persons developing asymptomatic/mild, severe, or critical infections was based on the modeled SARS-CoV-2 epidemic in France [20]. Severe and critical infections progress to severe and critical disease, respectively, prior to recovery. Patients are hospitalized in acute-care and ICU-care beds, respectively, based on existing standards of care in Qatar. Critical disease cases have an additional risk of COVID-19 mortality.

The model was parameterized using the best available data for SARS-CoV-2 natural history and epidemiology. A detailed description of model parameters, definitions, values, and justifications is found in Tables S1-S2 in the SM. The size and demographic structure of the population of Qatar were based on a population census conducted by Qatar’s Planning and Statistics Authority [5]. Life expectancy was obtained from the United Nations World Population Prospects database [21].

### Model fitting and analyses

The model was fitted to the standardized and centralized databases of SARS-CoV-2 testing, infections, hospitalizations, and mortality [8], as well as to findings of ongoing epidemiologic studies [8,11,22,23]. Data included: 1) time-series of the number of polymerase chain reaction (PCR)-confirmed SARS-CoV-2 cases, 2) time-series of the SARS-CoV-2 testing PCR positivity rate in each national subpopulation, 3) time-series of the PCR positivity rate in symptomatic patients with suspected SARS-CoV-2 infection coming to primary healthcare centers, 4) time-series of the proportion of laboratory-confirmed SARS-CoV-2 cases aged >60 years, 5) time-series of new/daily hospital admissions in acute-care beds and ICU-care beds, 6) the proportion of acute-care cases subsequently transferred to ICUs, 7) time-series of hospital occupancy in acute-care and ICU-care beds, 8) the cumulative number of deaths (not time series, due to the relatively small number of deaths), 9) a community survey assessing active-infections using PCR, 10) age-distribution of antibody positivity [8,22,23], and 11) national subpopulation distribution of antibody positivity [8,22,23]. A nonlinear least-square data fitting method, based on the Nelder-Mead simplex algorithm, was used to conduct the model fitting [24].

Model fitting was used to estimate epidemiologic indicators such as incidence, prevalence, attack rate (proportion of the population ever infected), and *R*_0_, as well as to forecast acute-care and ICU-care hospital admissions and hospital bed occupancy. The model was further used to evaluate the impact of implemented social and physical distancing restrictions by comparing model projections of the actual epidemic to those in a counter-factual scenario in which such interventions were not enforced. Informed by global estimates of *R*_0_ in the early epidemic that ranged between 2-4 [25,26], the counter-factual scenario with no interventions was implemented assuming *R*_0_ = 3. The model was also used to predict the impact of different scenarios for easing of social and physical distancing restrictions.

### Uncertainty analysis

Five hundred simulation runs were conducted to determine the range of uncertainty attending model predictions. At each run, Latin Hypercube sampling was applied in selecting input parameter values [27,28] from pre-specified ranges that assume ±30% uncertainty around parameter point estimates. The model was then refitted to input data. The resulting distribution for each model prediction, based on the 500 runs, was used to derive the mean and 95% uncertainty interval (UI).

Mathematical modeling analyses were conducted in MATLAB R2019a (Boston/MA/USA) [29] whereas statistical analyses were performed in STATA/SE 16.1 (College Station, TX) [30].

## Results

The model fitted the various data sources (examples in Figures S2-S3). Figure 1 shows model predictions for evolution of SARS-CoV-2 incidence, cumulative incidence, active-infection prevalence, and attack rate in the total population. Peak incidence was estimated at 12,750 new infections on May 23, 2020 while peak prevalence was estimated at 3.2% on May 25, 2020. By October 15, 2020, an estimated 1,426,500 infections were projected to have occurred, for a proportion of the population infected of 50.8%. Also by October 15, 2020, the proportion of all infections that had actually been diagnosed and confirmed by PCR was estimated at 11.6%. *R*_0_ varied between 1.07-2.78 from March 1 to October 15, with the highest values reached well after the onset of easing of restrictions on June 15, 2020 (Figure S4A of SM).

**Figure 1.**
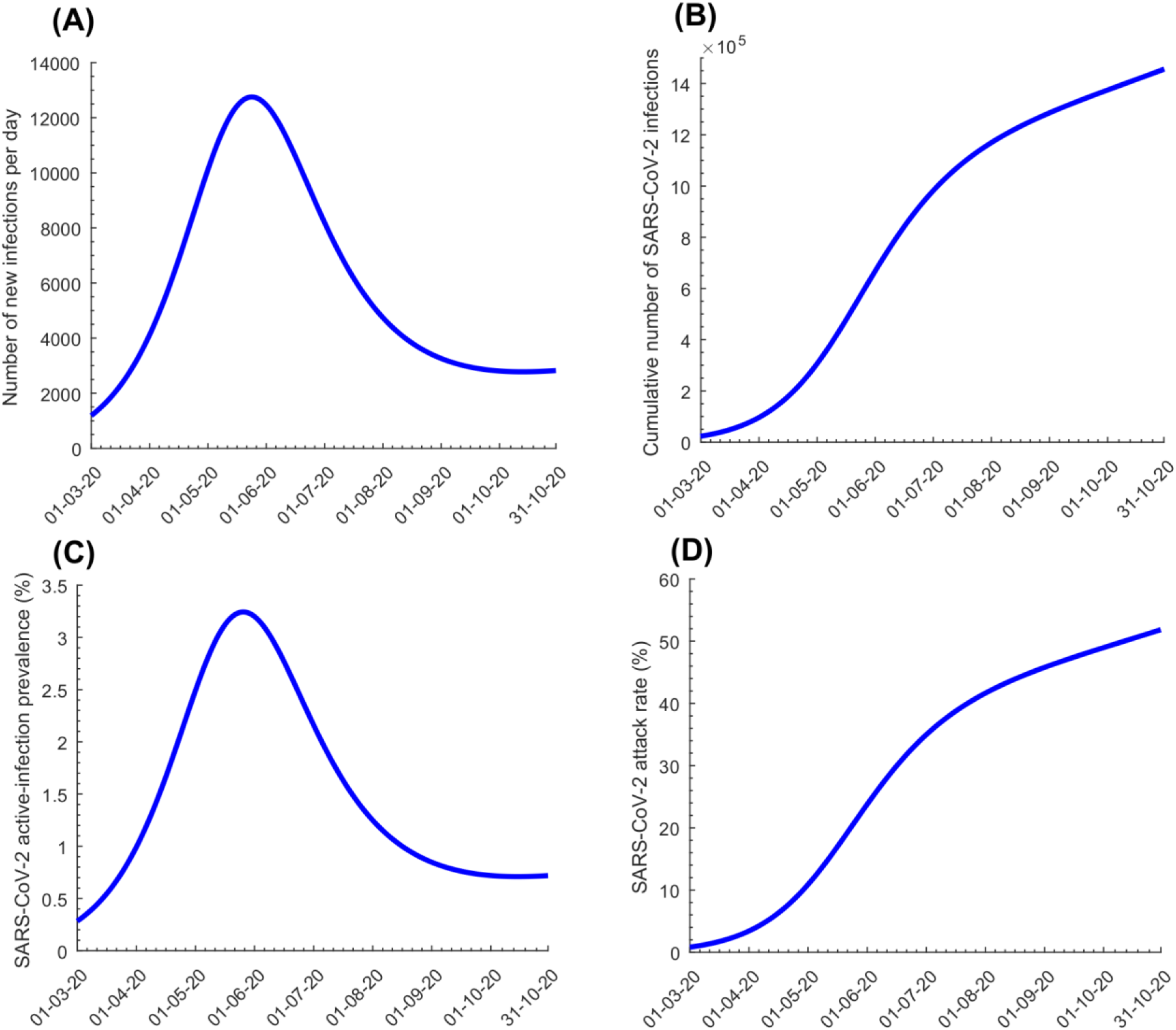
Model predictions for evolution of SARS-CoV-2 A) incidence (number of daily new infections), B) cumulative number of infections, C) active-infection prevalence (those latently infected or infectious), and D) attack rate (proportion ever infected) in the total population of Qatar.

Figure 2A-2B shows model-predicted daily hospital admissions in acute-care and ICU-care beds, respectively. New hospital admissions were predicted to peak at 292 acute-care beds on May 22, 2020 and 23 ICU-care beds on May 27, 2020. Figure 2C-2D shows evolution of hospital occupancy in acute-care and ICU-care beds. Peaks were predicted at 1,910 acute-care beds on May 27, 2020 and 244 ICU-care beds on June 6, 2020. The average hospital stay in an acute-care bed was estimated at 7.7 days while the stay in an ICU-care bed was estimated at 14.0 days. These model predictions agreed with actual COVID-19 hospital admission data (Figure S3 of SM).

**Figure 2.**
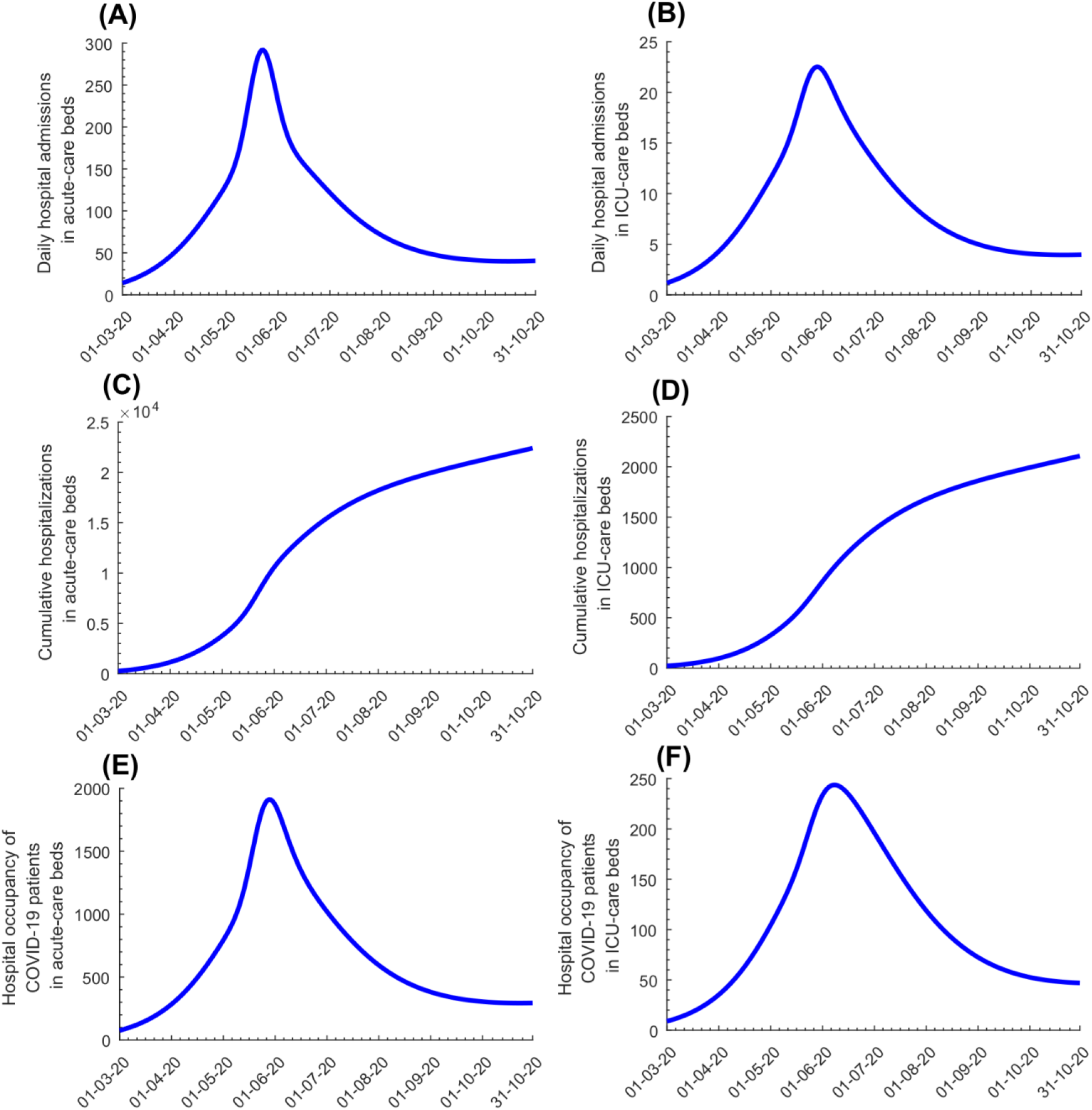
Model predictions for evolution of COVID-19 A) daily hospital admissions in acute-care beds, B) daily hospital admissions in ICU-care beds, C) cumulative number of hospitalizations in acute-care beds, D) cumulative number of hospitalizations in ICU-care beds, E) hospital occupancy of COVID-19 patients (number of beds occupied at any given time) in acute-care beds, and F) hospital occupancy of COVID-19 patients in ICU-care beds.

Discussions with policymakers to plan easing of social and physical distancing restrictions were initiated in April of 2020. The *effective* reproduction number (*R*_*t*_), the number of secondary infections each infection is generating at a given time, *t*, heavily influenced these discussions. Based on the model-predicted evolution of *R*_*t*_ *at that time* (Figure 3A), it was advised that no easing of restrictions should occur before the epidemic peak, then predicted to occur on May 20, as the epidemic was still in its exponential growth phase (*R*_*t*_ > 1). Model simulations confirmed that premature easing of restrictions would result in epidemic amplification (Figure 3B). To minimize the likelihood of a second wave and to buffer against a potential increased contact rate in the population, it was advised that easing of restrictions should not start before *R*_*t*_ reached 0.70, and that easing of restrictions should be implemented gradually over at least two months. Model simulations confirmed this rationale, and indicated that gradual easing of restrictions after *R*_*t*_ reached 0.70 would minimize the risk of a second wave (Figure 3C). Accordingly, policymakers planned and subsequently implemented a gradual easing of restrictions starting June 15, 2020, the day on which *R*_*t*_ was predicted to decline to 0.7. This line of analysis and rationale proved successful, as no second wave had materialized as of October 15, 2020, five months after the epidemic peak (Figure 3D).

**Figure 3.**
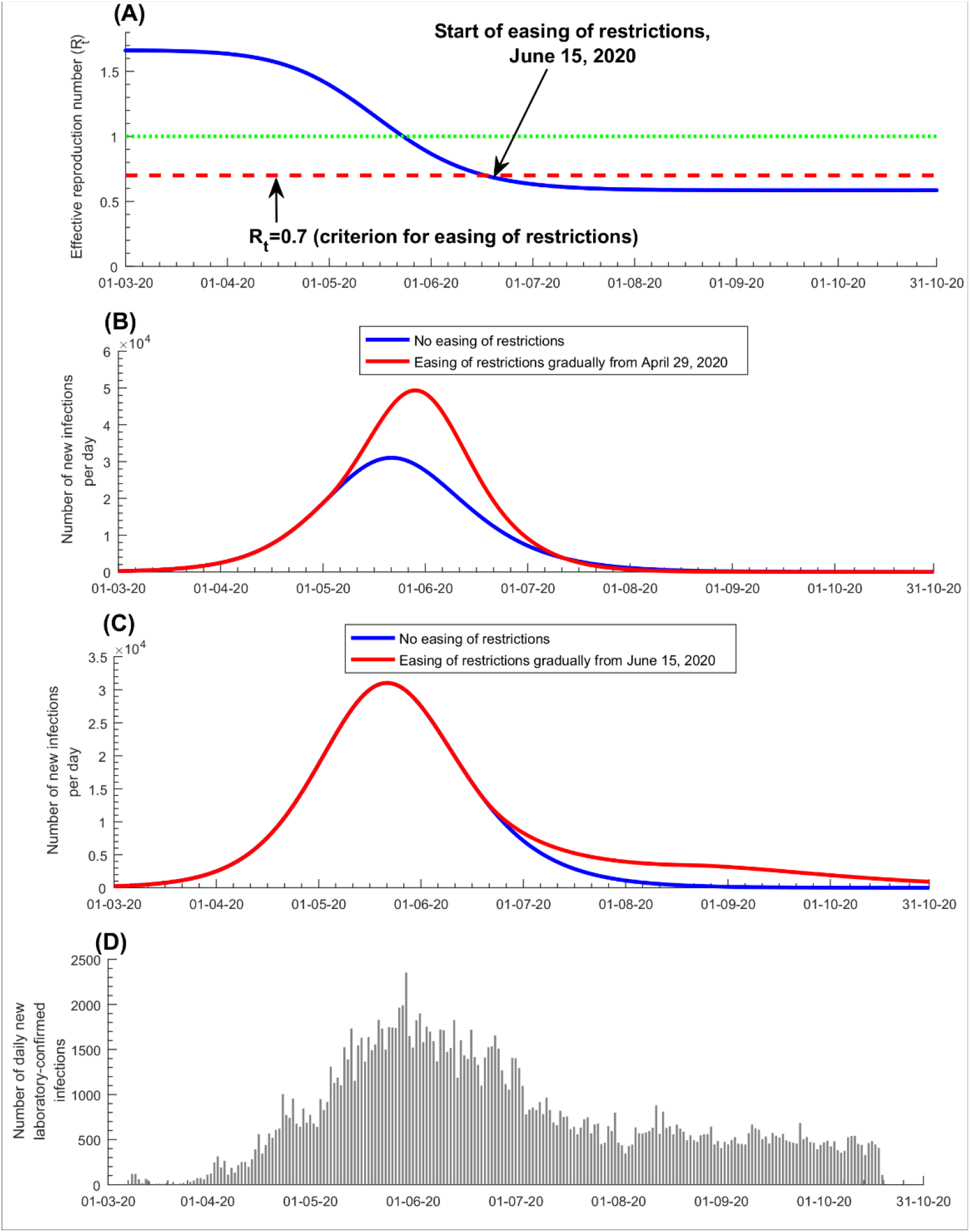
A) Effective reproduction number *R*_*t*_ and easing of social and physical distancing restrictions. B) Prediction of the number of daily new infections with early easing of restrictions, three weeks before the epidemic peak. C) Prediction of the number of daily new infections with delayed easing of restrictions, three weeks after the epidemic peak. This figure demonstrates the rationale and criteria used for the start of easing of restrictions. The figure shows the model fit and results *at the time* when the policy decision was actually made. An updated prediction for *R*_*t*_ is in Figure S4 of SM. The figure also shows in D) the number of daily new diagnosed and laboratory-confirmed infections.

Figures 4 and S5 (SM) show the predicted evolution of the epidemic in the *counter-factual scenario* of no social and physical distancing interventions. In the absence of these interventions, the epidemic would have peaked at 97,100 new infections per day on April 3, 2020 (Figure 4A), and at a prevalence of 23.4% on April 5, 2020 (Figure 4B). New hospital admissions would have peaked at 1,235 acute-care bed admissions on April 7, 2020 (Figure 4C) and at 103 ICU-care bed admissions on April 10, 2020 (Figure 4D). Accordingly, by October 15, 2020, the enforced social and physical distancing restrictions reduced the peaks for incidence, prevalence, and acute-care and ICU-care hospital admissions by >75% (Figure 4A-4D), and averted 840,000 infections (37%; Figure S5A of SM), 209 deaths (46%; Figure S5B of SM), 10,110 acute-care hospital admissions (32%; Figure S5C of SM), and 1,056 ICU-care hospital admissions (34%; Figure S5D of SM). These results show the extent of flattening of the epidemic curve that was accomplished with the enforced social and physical distancing interventions.

**Figure 4.**
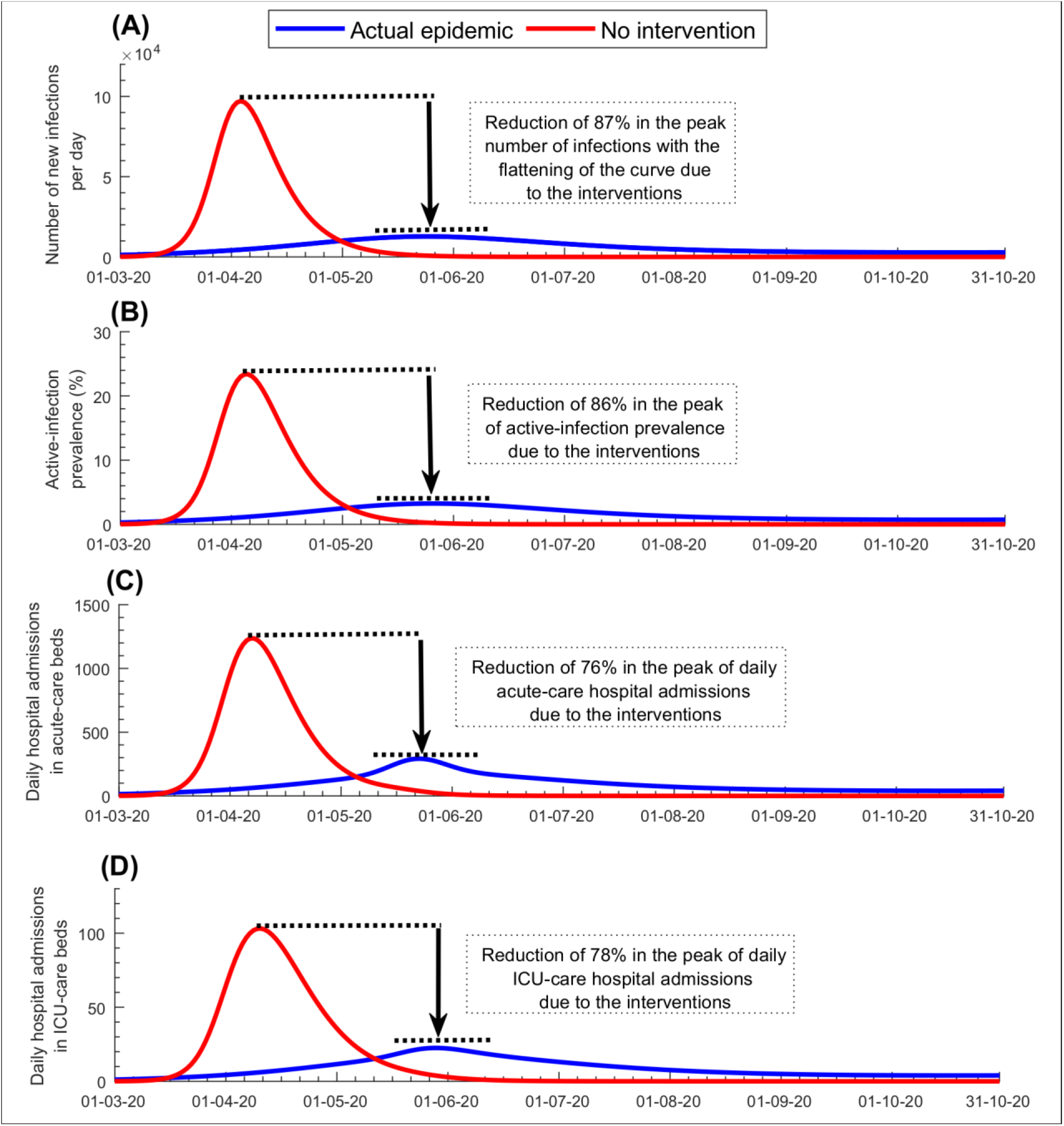
Impact of social and physical distancing interventions on A) number of daily new infections, B) active-infection prevalence (those latently infected or infectious), C) daily hospital admissions in acute-care beds, and D) daily hospital admissions in ICU-care beds.

Figure S6 of SM shows the results of the uncertainty analysis for the key epidemiological indicators in Figure 1, and Figure S7 of SM shows the results of the uncertainty analysis for the key hospitalization indicators in Figure 2. The results indicated overall narrow uncertainty intervals confirming the model’s predictive power.

## Discussion

Our study demonstrates that mathematical modeling was influential in informing the national public-health response and in formulating evidence-based policy decisions to minimize the pandemic’s toll on health, society, and the economy. The model, which was implemented in real-time, starting from March 2020, and was continuously updated and refined as more data became available, predicted with reasonable accuracy and precision the key epidemiologic indicators, such as the epidemic peak and the impact of easing of restrictions, as well as healthcare needs, at a time of uncertainty in which knowledge of the epidemiology of this infection was growing but still limited.

One of the highlights of this modeling approach is the application of the concept of *rational R*_*t*_ *tuning* for managing the easing of restrictions (Figure 4). Grounded on a theoretical foundation [4], rational *R*_*t*_ tuning proved to be a successful and effective strategy in safely easing the restrictions so as to ensure social and economic stability and functionality, while minimizing the risk of a second wave (Figure 3). Another highlight is the estimation of healthcare needs that guided resource-allocation planning well before the time when these resources were needed. Throughout the epidemic, including the epidemic peak, healthcare needs in Qatar remained well within the health system capacity, avoiding any serious strain. Importantly, this forecasting of healthcare needs also prevented resource waste by avoiding overestimation of healthcare needs. Despite the large number of infections in Qatar, results show that the epidemic would have been far worse if no social and physical distancing interventions had been enforced. In absence of interventions, the epidemic would have progressed very rapidly to a peak nearly 10-fold higher than what was actually observed (Figure 4). Disease burden would have been much larger and the healthcare system would have been strained to the point of collapse. This demonstrates that for a respiratory infection with such large *R*_0_ and serious disease sequalae, inaction would have had dire consequences, and that the national strategy focused on flattening the epidemic curve was appropriate to manage the epidemic.

An important finding of this study is that PCR-confirmed infections constitute only a small fraction of the actual number of infections. Only 11.6% of infections were estimated to have ever been diagnosed, probably because most infections were asymptomatic or mild. Indeed, a nation-wide population-based survey in Qatar showed that 58.5% of those who were PCR positive in this survey reported no symptoms during the last two weeks preceding the survey [8]. The growing number of serological testing studies in Qatar have also shown that the vast majority of those who are antibody-positive were never diagnosed with this infection [8,11,22,23]. For instance, out of all those antibody-positive in a nation-wide seroprevalence survey of the CMW population, only 9.3% had a documented, PCR-confirmed infection *prior* to antibody testing, affirming that as estimated by the model, nine of every 10 infections were never diagnosed. These findings are also consistent with a growing body of serological evidence from other countries [31-35].

We found that >97% of infections estimated to have occurred did not require hospitalization. The low infection severity appears to be a consequence of the young age profile of the population, with only 2% being >60 years of age [5,8,17,36], in addition to a well-funded healthcare system that emphasizes a proactive, high-quality standard of care [8], and possibly high levels of T cell cross-reactivity against SARS-CoV-2, reflecting T cell memory of circulating ‘common cold’ coronaviruses [37-41].

This study has limitations. Model estimates are contingent on the validity and generalizability of input data. Our estimates were based on current SARS-CoV-2 natural history and disease progression parameters, but our understanding of this infection is still evolving. Available input data were most complete at the national level. We did not have sufficient data about social networks of different national subpopulations and patterns of mixing between those subpopulations to factor them into the model. Despite these limitations, our model, tailored to the complexity of the epidemic in Qatar, was able to reproduce observed epidemic trends, and to provide useful and consequential predictions and insights about infection transmission and healthcare needs. Importantly, the modeling estimates successfully influenced the national response.

In conclusion, Qatar experienced a large SARS-CoV-2 epidemic, but avoided a burdensome epidemic, such as that unfolding in other counties. Mathematical modeling played an influential role in guiding the national public-health response by characterizing and understanding the epidemic, forecasting healthcare needs, predicting the impact of social and physical distancing restrictions, and rationalizing and justifying the easing of restrictions. While this article illustrates a successful case study, the modeling tools employed here can be adapted and applied in other countries to guide SARS-CoV-2 epidemic control, preparedness for the current or future waves of infection, or enforcement and easing of restrictions or other interventions, such as vaccination [19].

## Data Availability

All data are available within the manuscript and its supplementary material.

## Acknowledgements

We thank Her Excellency Dr. Hanan Al Kuwari, Minister of Public Health, for her vision, guidance, leadership, and support. We also thank Dr. Saad Al Kaabi, Chair of the System Wide Incident Command and Control (SWICC) Committee for the COVID-19 national healthcare response, for his leadership, analytical insights, and for his instrumental role in enacting data information systems that made these studies possible. We further extend our appreciation to the SWICC Committee and the Scientific Reference and Research Taskforce (SRRT) members for their informative input, scientific technical advice, and enriching discussions. We also thank Dr. Mariam Abdulmalik, CEO of the Primary Health Care Corporation and the Chairperson of the Tactical Community Command Group on COVID-19, as well as members of this committee, for providing support to the teams that worked on the field surveillance. We also acknowledge the dedicated efforts of the Clinical Coding Team and the COVID-19 Mortality Review Team, both at Hamad Medical Corporation, and the Surveillance Team at the Ministry of Public Health.

## Funding

This work was supported by the Biomedical Research Program and the Biostatistics, Epidemiology and Biomathematics Research Core, both at Weill Cornell Medicine-Qatar, and by the Ministry of Public Health and Hamad Medical Corporation. Statements made herein are solely the responsibility of the authors.

## Author contributions

HHA co-designed the study, constructed and parameterized the mathematical model and conducted the mathematical modeling analyses. HC conducted the statistical analyses, contributed to the parameterization of the mathematical model, and wrote the first draft of the manuscript. LJA conceived and co-designed the study and led the construct and parameterization of the mathematical model, conduct of analyses, and drafting of the article. All authors contributed to conceptualization of the analyses, discussion and interpretation of the results, and writing of the manuscript. All authors have read and approved the final manuscript.

## Competing interests

We declare no competing interests.

## Supplementary Material

### I., SARS-CoV-2 mathematical model structure

A deterministic age-structured meta-population compartmental model was developed to describe the severe acute respiratory syndrome coronavirus 2 (SARS-CoV-2) transmission dynamics and disease progression in the population of Qatar, factoring subpopulation heterogeneity in exposure to the infection. The model stratified the population into compartments according to nationality subpopulation, age group (0-9, 10-19, 20-29,…, ≥80 years), infection status (infected, uninfected), infection stage (asymptomatic/mild, severe, critical), and disease stage (severe disease or critical disease). All Coronavirus Disease 2019 (COVID-19) mortality was assumed to occur in individuals that are in the critical disease stage. The model is based on extension and adaptation of our calibrated mathematical models developed to characterize SARS-CoV-2 transmission dynamics [1-5].

Epidemic dynamics were described using a system of coupled nonlinear differential equations for each age group and subpopulation (nationality) group. Each age group, *a*, denoted a ten-year age band apart from the last category which grouped together all individuals ≥80 years of age. The population was divided into seven resident subpopulation groups *i* (*i* = 1, 2, 3, 4, 5, 6, 7) representing the subpopulations of Indians, Bangladeshis, Nepalese, Qataris, Egyptians, Filipinos, and all other nationalities, respectively—these are the largest nationality subpopulation groups in Qatar. Qatar’s population composition and subpopulations size and demographic structure were based on findings of “The Simplified Census of Population, Housing, and Establishments” conducted by Qatar’s Planning and Statistics Authority [6]. Life expectancy was obtained from the United Nations World Population Prospects database [7].

**Figure S1.**
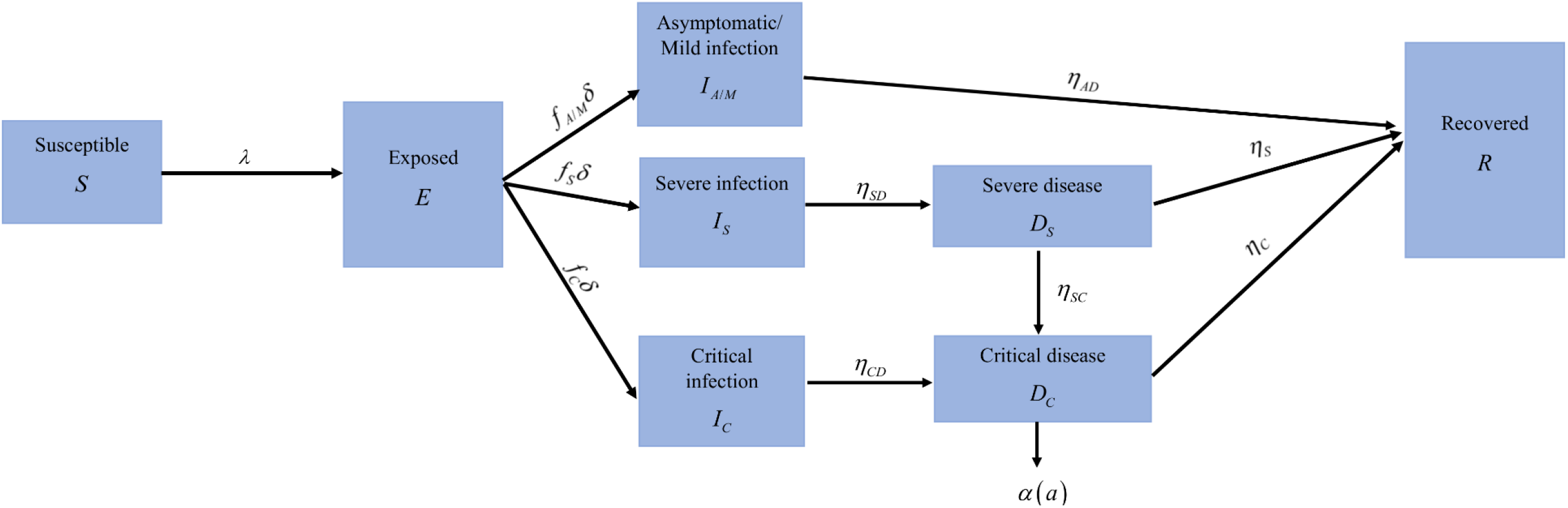
Schematic diagram describing the basic structure of the SARS-CoV-2 mathematical model.

The model was expressed in terms of the following system of coupled nonlinear differential equations for each subpopulation group and age group:

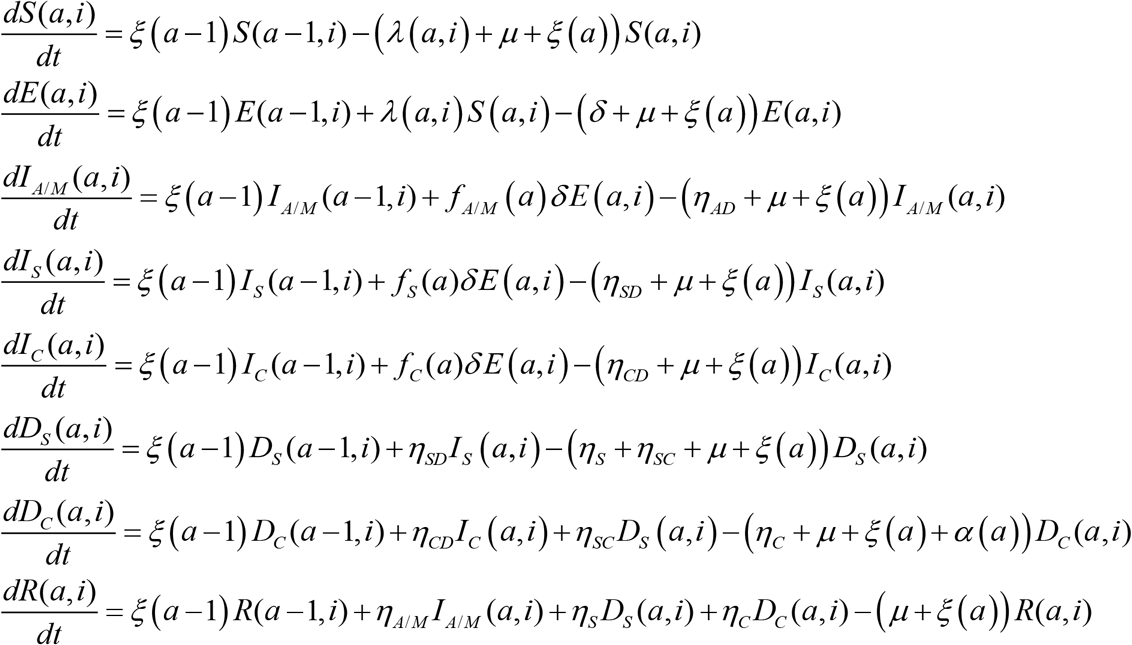

The definitions of population variables and symbols used in the equations are in Table S1.

**Table S1.** Definitions of population variables and symbols used in the model

| Symbol | Definition |
| --- | --- |
| $S(a,i)$ | Susceptible population |
| $E(a,i)$ | Latently infected population |
| $I_{A/M}(a,i)$ | Population with asymptomatic/mild infection |
| $I_S(a,i)$ | Population with severe infection |
| $I_C(a,i)$ | Population with critical infection |
| $D_S(a,i)$ | Population with hospitalization in acute-care beds |
| $D_C(a,i)$ | Population with hospitalization in intensive care unit beds |
| $R(a,i)$ | Recovered population |
| $n_{age}$ | Number of age groups |
| $n_{pop}$ | Number of subpopulation groups |
| $\xi(a)$ | Transition rate from one age group to the next age group. Here $\xi(0) = \xi(n_{age}) = 0$ |
| $\sigma(a)$ | Susceptibility profile to the infection in each age group |
| $\psi(i)$ | Level of exposure profile in each subpopulation group $i$ |
| $1/\delta$ | Duration of latent infection |
| $\beta$ | Average rate of infectious contacts |
| $1/\eta_{AD}$ | Duration of asymptomatic/mild infection infectiousness |
| $1/\eta_{SD}$ | Duration of severe infection infectiousness before isolation and/or hospitalization |
| $1/\eta_S$ | Duration of severe disease following onset of severe disease |
| $1/\eta_{CD}$ | Duration of critical infection infectiousness before isolation and/or hospitalization |
| $1/\eta_C$ | Duration of critical disease following onset of critical disease |
| $1/\eta_{SC}$ | Rate of disease progression from severe disease to critical disease |
| $\alpha(a)$ | Disease mortality rate in each age group |
| $1/\mu$ | Natural death rate |
| $f_{A/M}(a)$ | Proportion of infections that will progress to be asymptomatic/mild infections |
| $f_S(a)$ | Proportion of infections that will progress to be infections that require hospitalization in acute-care beds |
| $f_C(a)$ | Proportion of infections that will progress to be infections that require hospitalization in intensive care unit beds |
| $Z$ | The subpopulation mixing matrix |
| $H$ | The age-mixing matrix |

The force of infection *λ*(*a,i*) (hazard rate of infection) experienced by each susceptible *S*(*a,i*) population, is given by

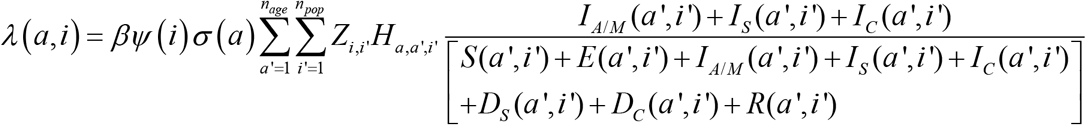

Here *β* is the rate of infectious contacts, *Ψ* (*i*) is the level of exposure profile in each subpopulation group *i*, and *σ* (*a*) is the susceptibility profile to the infection in each age group *a*.

To account for temporal variation in the basic reproduction number (*R*_0_), we incorporated temporal changes in the rate of infectious contacts. We parameterized the temporal variation (time dependence of *β*) through the following combined function of the Woods-Saxon and logistic functions.

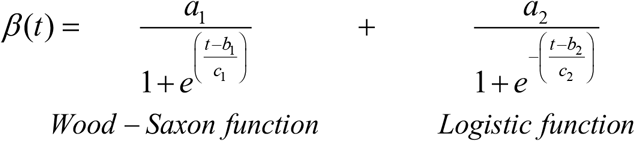

This function was mathematically designed to describe and characterize the time evolution of the level of risk of exposure before and after easing of restrictions. It was informed by our knowledge of SARS-CoV-2 epidemiology in Qatar [4], and it provided a robust fit to the data. Here *a*_1_, *a*_2_, *b*_1_, *b*_2_, *c*_1_, and *c*_2_ are fitting parameters.

The mixing among the different age groups and subpopulation groups is dictated by the mixing matrices *H*_*a,a* ′,*i* ′_ (for age group mixing) and *Z*_*i,i′*_ (for subpopulation group mixing). These matrices provide the likelihood of mixing and are given by the following expressions:

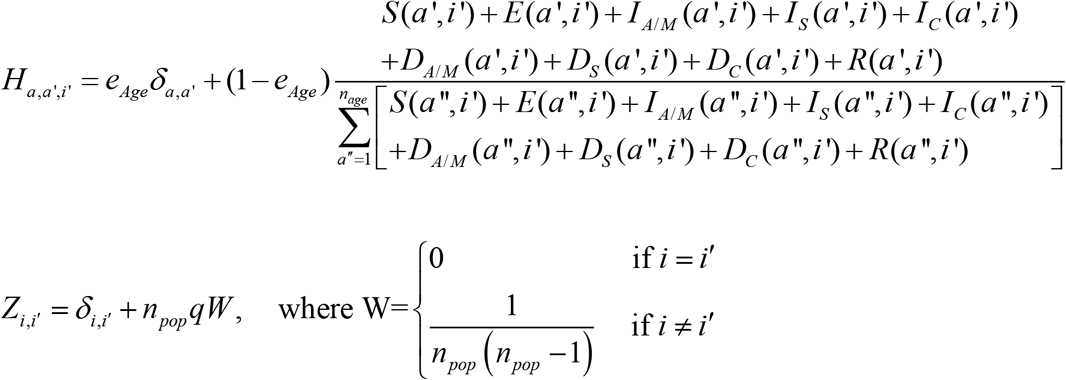

Here, *δ*_*a,a ′*_ (and *δ*_*i,i′*_) is the identity matrix. *e*_*Age*_ ∈[0,1] measures the degree of assortativeness in the age mixing. At the extreme *e*_*Age*_ = 0, the mixing is fully proportional, while at the other extreme, *e*_*Age*_ = 1, the mixing is fully assortative, that is individuals mix only with members in their own age group. *W* is the subpopulation connectivity matrix of dimension *n*_*pop*_ × *n*_*pop*_ and the mixing is assumed symmetric for all subpopulations. The *W* matrix normalizes to 1, that is the sum of all entries adds up to 1. *q* parametrizes the connectivity to other subpopulations relative to the connectivity within the same subpopulation.

### II., Model fitting and parameter values

The model was fitted to the following sources of data: 1) time-series of the number of polymerase chain reaction (PCR) laboratory-confirmed SARS-CoV-2 cases, 2) time-series of SARS-CoV-2 testing PCR positivity rate in each nationality subpopulation, 3) time-series of PCR positivity rate in symptomatic patients with suspected SARS-CoV-2 infection presenting to primary healthcare centers, 4) time-series of proportion of laboratory-confirmed SARS-CoV-2 cases aged >60 years, 5) time-series of new/daily hospital admissions in acute-care beds and in ICU-care beds, 6) proportion of acute-care bed cases transferred subsequently to ICU-care beds, 7) time-series of hospital occupancy in acute-care beds and in ICU-care beds, 8) cumulative number of deaths (not time series with the relatively small number of deaths), 9) one community survey assessing active-infection using PCR, 10) age-distribution of antibody positivity [4,8,9], and 11) nationality subpopulation distribution of antibody positivity [4,8,9].

Model input parameters were based on best available empirical data for SARS-CoV-2 natural history and epidemiology. Model parameter values are listed in Table 2. The following parameters were derived by fitting the model to data: *f*_*C*_, *η*_*S*_, *η*_*C*_, *η*_*SC*_, *α, σ* (*a*), *Ψ* (*i*), *e*_*Age*_, *q, a*_0_, *a*_1_, *a*_2_, *b*_0_, *b*_1_, *b*_2_, *t*_0_, *c*_1_, and *c*_2_.

**Table S2.** Model parameter values.

| Parameter | Symbol | Value | Justification |
| --- | --- | --- | --- |
| Duration of latent infection | $1/\delta$ | 3.69 days | Based on existing estimate [10] and based on a median incubation period of 5.1 days [11] adjusted by observed viral load among infected persons [12] and reported transmission before onset of symptoms [13]. |
| Duration of infectiousness | $1/\eta_{AD}$<br>$1/\eta_{SD}$<br>$1/\eta_{CD}$ | 3.48 days | Based on existing estimate[10] and based on observed time to recovery among persons with mild infection [10,14] and observed viral load in infected persons [12,13,15]. |
| Life expectancy in Qatar | $1/\mu$ | 80.7 years | United Nations World Population Prospects database [7]. |
| Disease mortality rate in each age group | $\alpha(a)$ | | The distribution and age dependence of COVID-19 mortality was based on the modeled SARS-CoV-2 epidemic in France [16]. |
| Age 0-19 years | $RRD1 \times \alpha$ | $RRD1 = 0.1$ | Model-estimated relative risk of death based on the SARS-CoV-2 epidemic in France [16]. |
| Age 20-29 years | $RRD2 \times \alpha$ | $RRD2 = 0.4$ | Model-estimated relative risk of death based on the SARS-CoV-2 epidemic in France [16]. |
| Age 30-39 years | $\alpha$ | Reference category | Model fitting |
| Age 40-49 years | $RRD3 \times \alpha$ | $RRD3 = 3.0$ | Model-estimated relative risk of death based on the SARS-CoV-2 epidemic in France [16]. |
| Age 50-59 years | $RRD4 \times \alpha$ | $RRD4 = 10.0$ | Model-estimated relative risk of death based on the SARS-CoV-2 epidemic in France [16]. |
| Age 60-69 years | $RRD5 \times \alpha$ | $RRD5 = 45.0$ | Model-estimated relative risk of death based on the SARS-CoV-2 epidemic in France [16]. |
| Age 70-79 years | $RRD6 \times \alpha$ | $RRD6 = 120.0$ | Model-estimated relative risk of death based on the SARS-CoV-2 epidemic in France [16]. |
| Age 80+ years | $RRD7 \times \alpha$ | $RRD7 = 505.0$ | Model-estimated relative risk of death based on the SARS-CoV-2 epidemic in France [16]. |
| Proportion of infections that will progress to be infections that require hospitalization in acute-care beds | $f_s(a)$ | | The distribution and age dependence of asymptomatic/mild, severe, or critical infections was based on the modeled SARS-CoV-2 epidemic in France [16]. |
| Age 0-19 years | $RRS1 \times f_s(t)$ | $RRS1 = 0.1$ | Model-estimated relative risk of severe infection based on the SARS-CoV-2 epidemic in France [16]. |
| Age 20-29 years | $RRS2 \times f_s(t)$ | $RRS2 = 0.5$ | Model-estimated relative risk of severe infection based on the SARS-CoV-2 epidemic in France [16]. |
| Age 30-39 years | $f_s(t) = a_0 e^{-\left(\frac{t-t_0}{b_0}\right)^2}$ | Reference category | This parameter was described by a Gaussian function, as it provided the best fit for the data of the daily hospital admissions in acute-care beds. |
| Age 40-49 years | $RRS3 \times f_s(t)$ | $RRS3 = 1.2$ | Model-estimated relative risk of severe infection based on the SARS-CoV-2 epidemic in France [16]. |
| Age 50-59 years | $RRS4 \times f_s(t)$ | $RRS4 = 2.3$ | Model-estimated relative risk of severe infection based on the SARS-CoV-2 epidemic in France [16]. |
| Age 60-69 years | $RRS5 \times f_s(t)$ | $RRS5 = 4.5$ | Model-estimated relative risk of severe infection based on the SARS-CoV-2 epidemic in France [16]. |
| Age 70-79 years | $RRS6 \times f_s(t)$ | $RRS6 = 7.8$ | Model-estimated relative risk of severe infection based on the SARS-CoV-2 epidemic in France [16]. |
| Age 80+ years | $RRS7 \times f_s(t)$ | $RRS7 = 27.6$ | Model-estimated relative risk of severe infection based on the SARS-CoV-2 epidemic in France [16]. |
| Proportion of infections that will progress to be infections that require hospitalization in intensive care unit beds | $f_c(a)$ | | The distribution and age dependence of asymptomatic/mild, severe, or critical infections was based on the modeled SARS-CoV-2 epidemic in France [16]. |
| Age 0-19 years | $RRC1 \times f_c$ | $RRC1 = 0.2$ | Model-estimated relative risk of critical infection based on the SARS-CoV-2 epidemic in France [16]. |
| Age 20-29 years | $RRC2 \times f_c$ | $RRC2 = 0.3$ | Model-estimated relative risk of critical infection based on the SARS-CoV-2 epidemic in France [16]. |
| Age 30-39 years | $f_c$ | Reference category | Model fitting |
| Age 40-49 years | $RRC3 \times f_c$ | $RRC3 = 1.8$ | Model-estimated relative risk of critical infection based on the SARS-CoV-2 epidemic in France [16]. |
| Age 50-59 years | $RRC4 \times f_c$ | $RRC4 = 4.7$ | Model-estimated relative risk of critical infection based on the SARS-CoV-2 epidemic in France [16]. |
| Age 60-69 years | $RRC5 \times f_c$ | $RRC5 = 10.6$ | Model-estimated relative risk of critical infection based on the SARS-CoV-2 epidemic in France [16]. |
| Age 70-79 years | $RRC6 \times f_c$ | $RRC6 = 13.6$ | Model-estimated relative risk of critical infection based on the SARS-CoV-2 epidemic in France [16]. |
| Age 80+ years | $RRC7 \times f_c$ | $RRC7 = 8.7$ | Model-estimated relative risk of critical infection based on the SARS-CoV-2 epidemic in France [16]. |

### III., Basic reproduction number *R*_0_ and effective reproduction number *R*_*t*_

As informed by the method of Heffernan and *et al*. [17], the overall basic reproduction number (*R*_0_) and overall effective reproduction number (*R*_*t*_) were derived to be

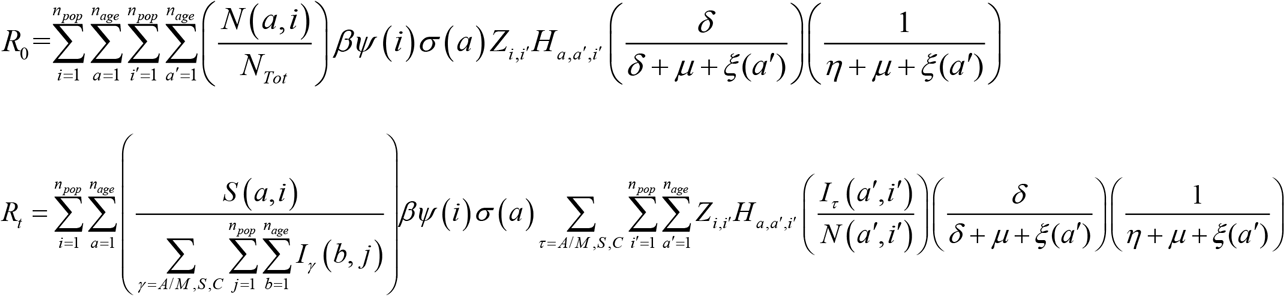

Here,*N*_*Tot*_ is the total population size and *N* (*a, i*) is the population size of each age group *a* and subpopulation group *i*.

**Figure S2.**
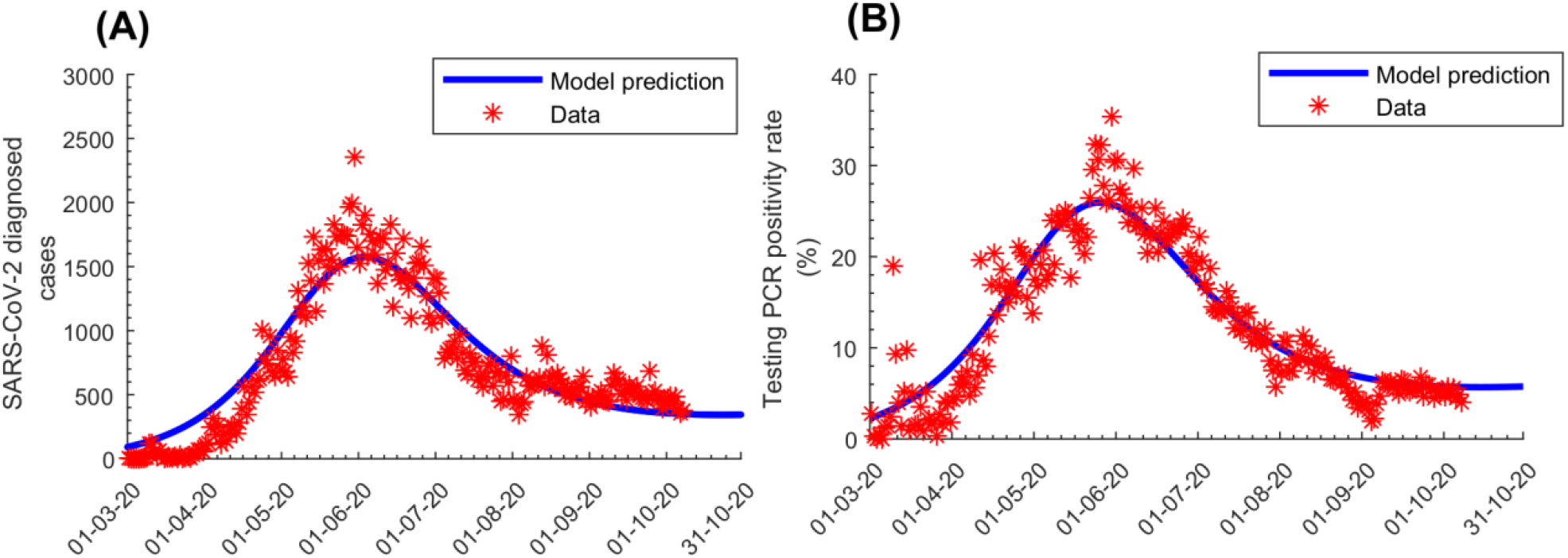
Model fits to (A) SARS-CoV-2 laboratory-confirmed cases and (B) testing PCR positivity rate.

**Figure S3.**
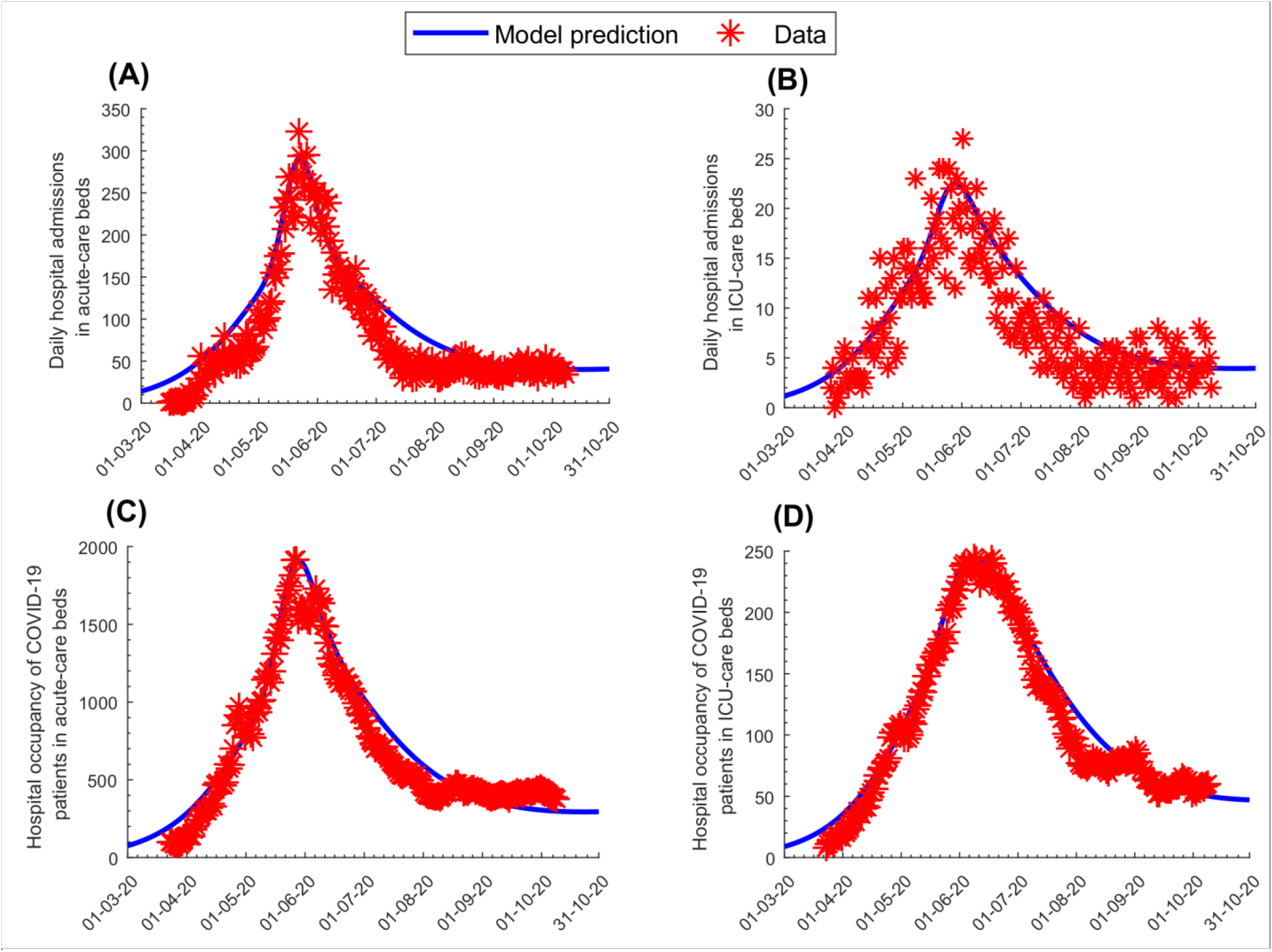
Model fits to A) daily hospital admissions in acute-care beds, B) daily hospital admissions in ICU-care beds, C) hospital occupancy of COVID-19 patients (number of beds occupied at any given time) in acute-care beds, and D) hospital occupancy of COVID-19 patients in ICU-care beds.

**Figure S4.**
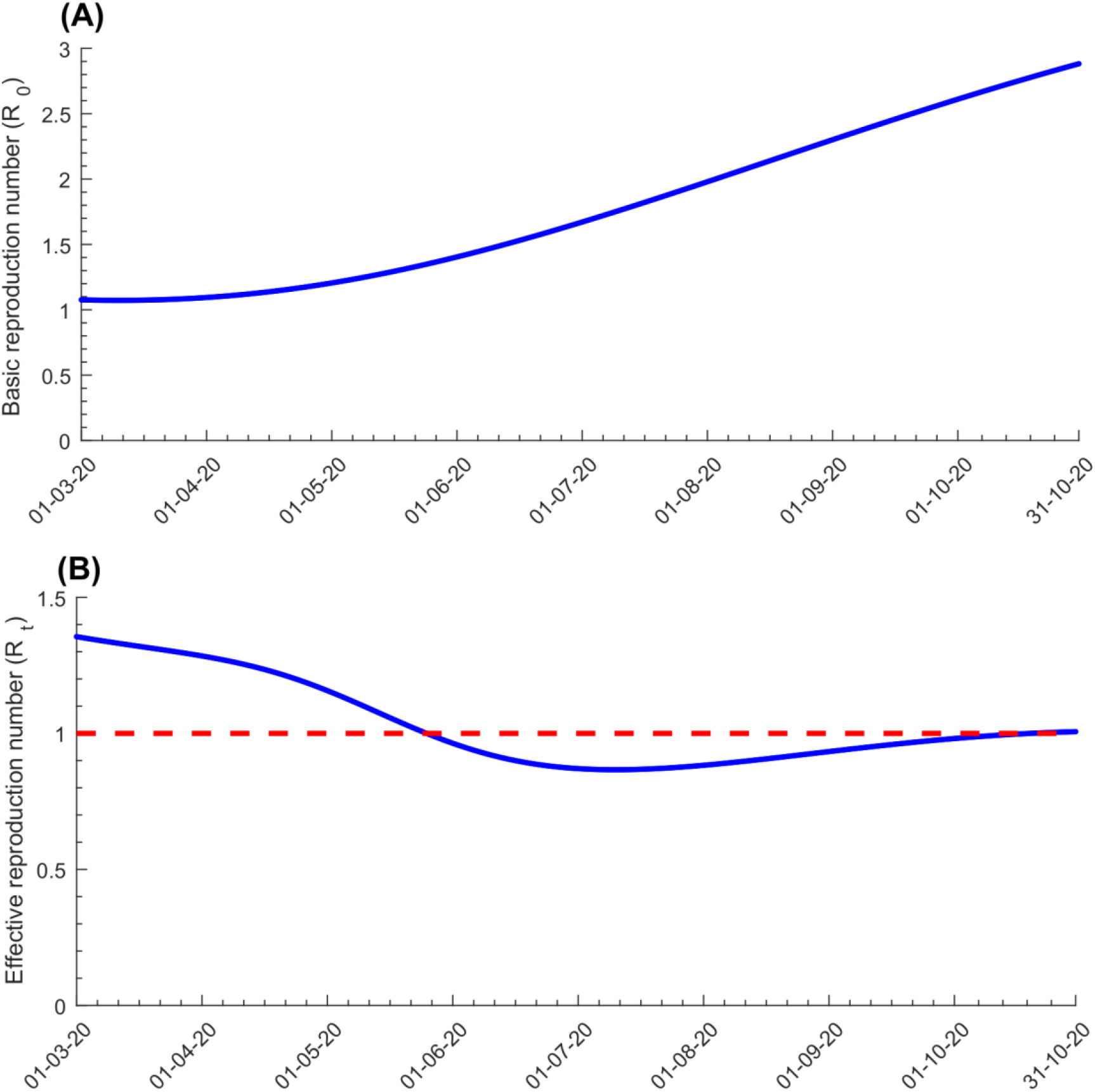
Evolution of the basic reproduction number *R*_*0*_ (A) and effective reproduction number *R*_*t*_ (B) in Qatar.

**Figure S5.**
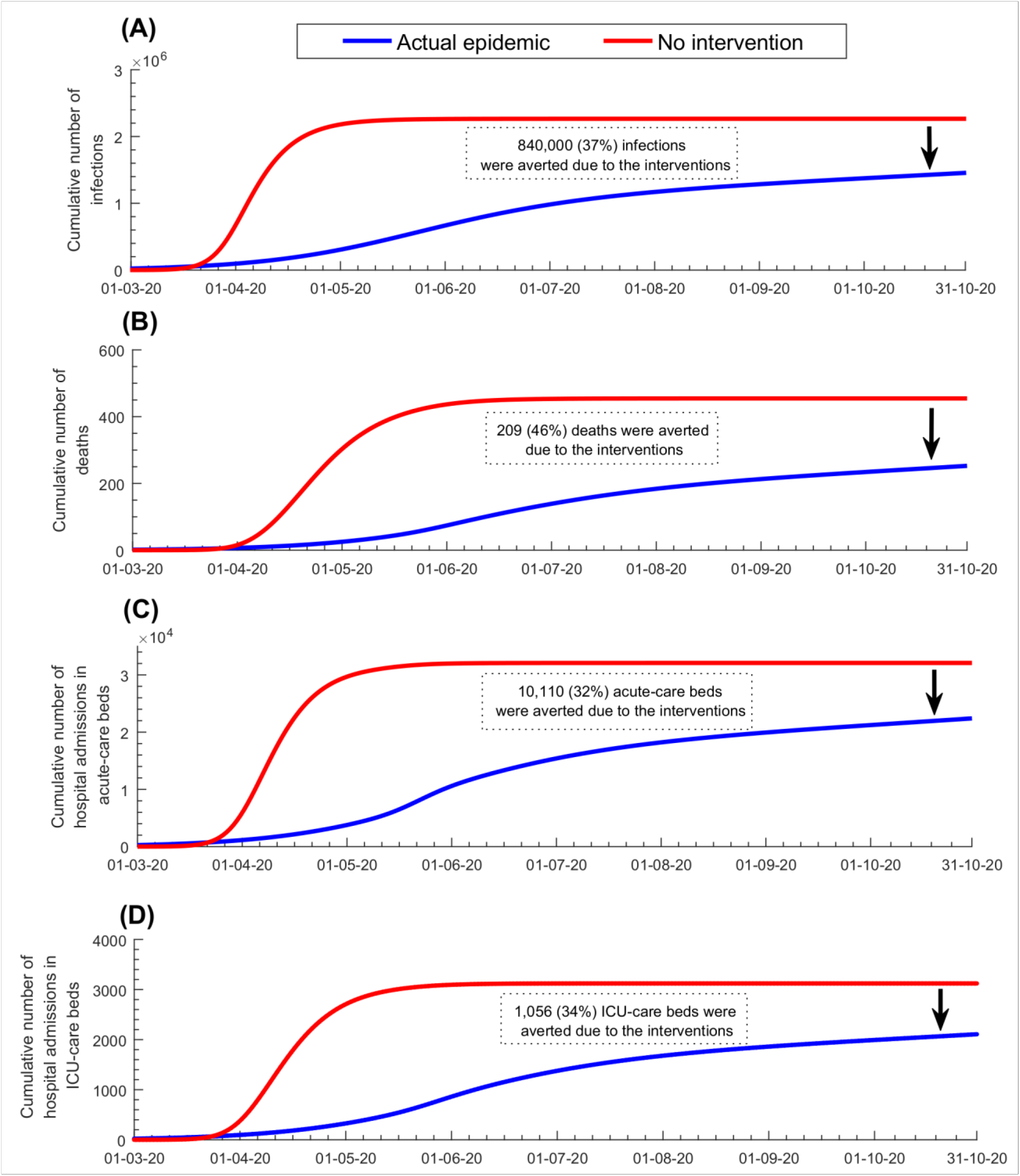
Impact of the social and physical distancing interventions on A) cumulative number of infections, B) cumulative number of deaths, C) cumulative number of hospital admissions in acute-care beds, and D) cumulative number of hospital admissions in ICU-care beds.

**Figure S6.**
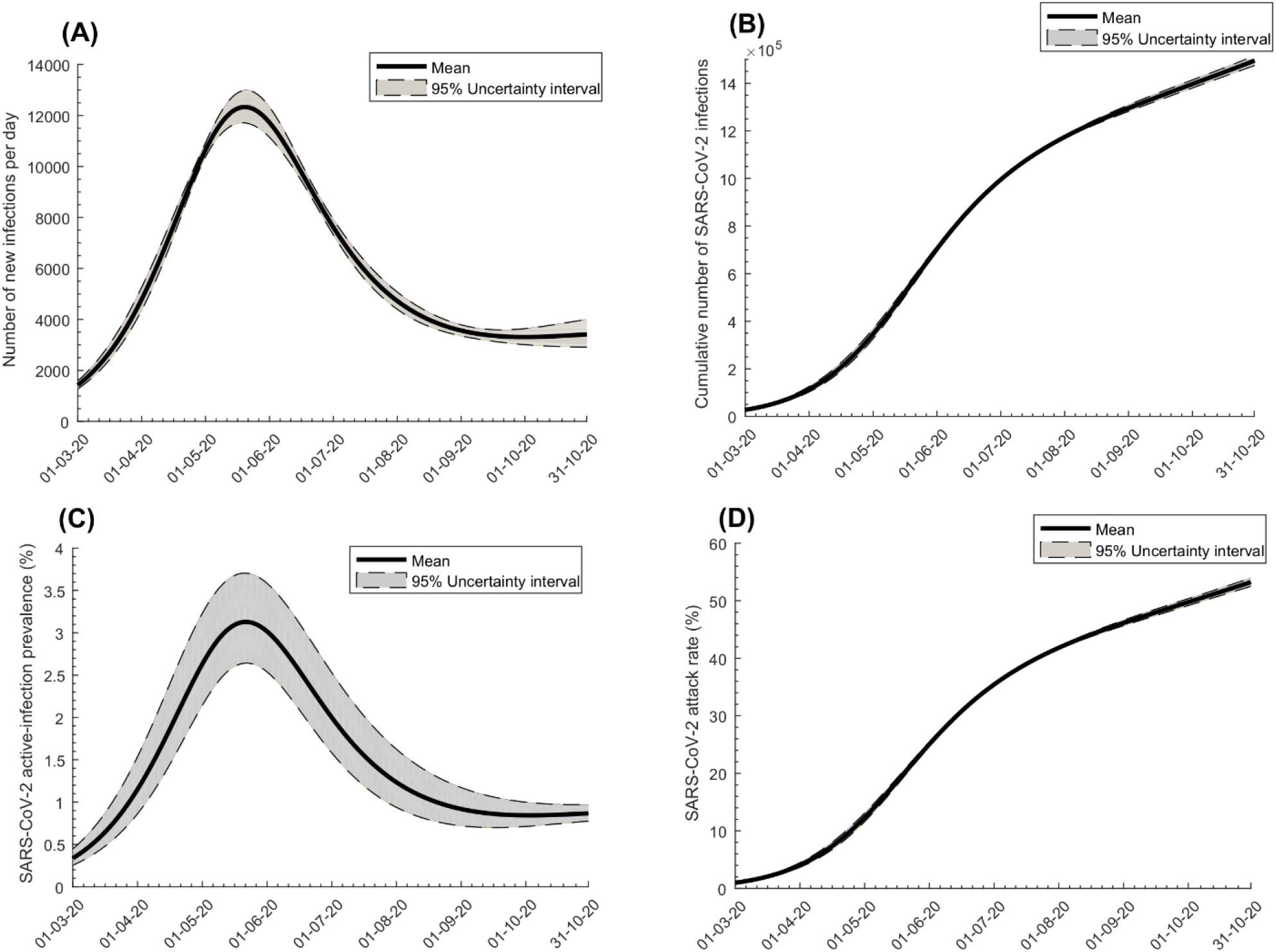
Uncertainty analysis. Mean and 95% uncertainty interval (UI) for the evolution of SARS-CoV-2 A) incidence (number of daily new infections), B) cumulative number of infections, C) active-infection prevalence (those latently infected or infectious), and D) attack rate (proportion ever infected), in the total population of Qatar.

**Figure S7.**
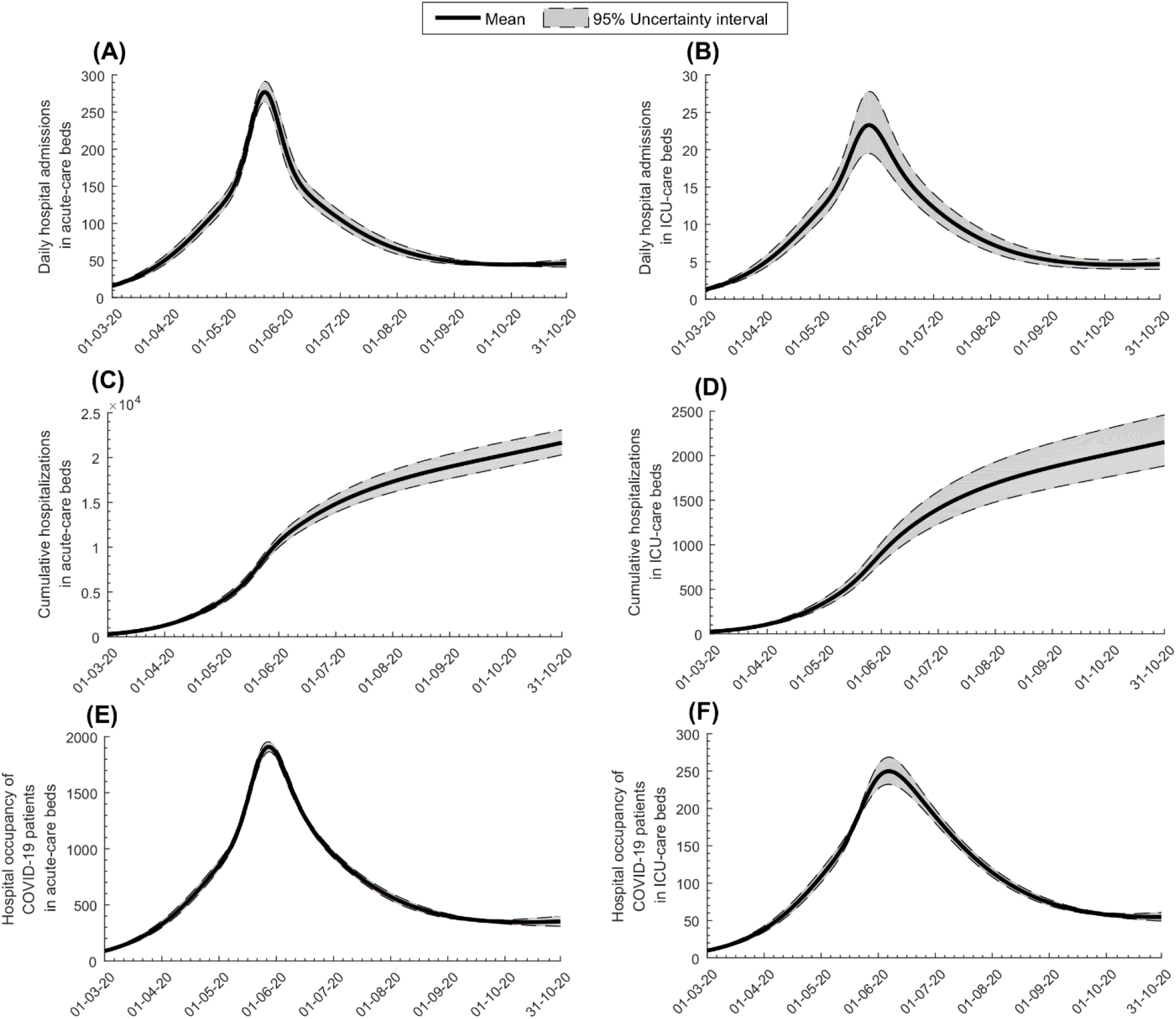
Uncertainty analysis. Mean and 95% uncertainty interval (UI) for the evolution of COVID-19 A) daily hospital admissions in acute-care beds, B) daily hospital admissions in ICU-care beds, C) cumulative number of hospitalizations in acute-care beds, D) cumulative number of hospitalizations in ICU-care beds, E) hospital occupancy of COVID-19 patients (number of beds occupied at any given time) in acute-care beds, and F) hospital occupancy of COVID-19 patients in ICU-care beds.

## Notes

### Competing Interest Statement

The authors have declared no competing interest.

